# Enhancing Diagnostic Precision in Thyroid Nodule Classification: A Deep Learning Approach to Automated Ultrasound Image Analysis

**DOI:** 10.1101/2025.02.05.25321737

**Authors:** Luís Jesuino de Oliveira Andrade, Gabriela Correia Matos de Oliveira, João Cláudio Nunes Carneiro Andrade, Alcina Maria Vinhaes Bittencourt, Adriana Malta de Figueiredo, Luís Matos de Oliveira

## Abstract

**Introduction:** Escalating thyroid nodule prevalence necessitates precise ultrasonographic diagnosis, which is constrained by operator-dependent variability. Convolutional neural network (CNN)-based artificial intelligence (AI)/machine learning (ML) frameworks can improve segmentation, malignancy prediction, and interobserver concordance, yet they often lack real-world clinical validation, interpretable architectures, and actionable validation frameworks for translational integration.

**Objective:** To improve diagnostic accuracy in thyroid nodule classification using a deep learning (DL) approach for automated analysis of ultrasound images.

**Method:** This methodology employed a multicenter, retrospective cohort of anonymized thyroid ultrasound images (benign/malignant, histopathology-confirmed) sourced from PubMed®. Images were preprocessed (normalization, denoising) with expert-annotated regions of interest (ROIs). A CNN-based DL framework (ResNet-50, EfficientNet-B0) was fine-tuned via transfer learning for automated nodule detection, segmentation, and malignancy classification aligned with ACR TI-RADS™ criteria. Validation utilized an independent test set, diagnostic metrics (sensitivity, specificity, area under the receiver operating characteristic curve (AUC-ROC)), and interobserver analysis (Cohen’s kappa) against three sonographers. Statistical rigor included PSPP-driven paired t-tests, chi-square tests, and McNemar’s tests to quantify AI-human concordance and optimize ACR TI-RADS™ integration for risk stratification.

**Results:** The AI model demonstrated high diagnostic efficacy: sensitivity 92.5%, specificity 88.3%, accuracy 90.4%, and AUC-ROC 0.94, surpassing sonographers in both sensitivity (p<0.001) and specificity (p<0.01). Interobserver concordance (Cohen’s κ=0.89) exceeded human variability (κ=0.72–0.85). ACR TI-RADS™ integration achieved 91.2% agreement, enhancing objectivity in the assessment of intermediate-risk nodules (categories 3–4). Feature analysis highlighted robust detection of hypoechoic patterns (94.2% sensitivity) and irregular margins (91.8% sensitivity), aligning with ACR TI-RADS™ criteria and confirming the AI’s potential to standardize risk stratification and reduce diagnostic subjectivity.

**Conclusion:** Advanced AI enhances thyroid ultrasound diagnostics through precise nodule detection and classification, reduced interobserver variability, and ACR TI-RADS™-aligned feature extraction, thereby boosting diagnostic confidence and clinical decision-making.

## INTRODUCTION

Thyroid cancer has emerged as one of the most prevalent endocrine malignancies worldwide, underscoring the importance of early and accurate diagnosis for effective treatment and improved patient outcomes.^1^ Thyroid ultrasonography has become a fundamental examination in thyroid imaging, offering real-time visualization and guiding fine-needle aspiration biopsies (FNAB).^2^ However, its diagnostic accuracy is often limited by subjective interpretation, variability in operator expertise, and the inherent challenges of distinguishing malignant from benign nodules.^2^ These limitations highlight the urgent need for innovative approaches to enhance the precision of thyroid ultrasound diagnostics.

Artificial intelligence (AI), particularly through machine learning (ML) and deep learning (DL), has revolutionized medical imaging by enabling the analysis of complex datasets and identifying patterns imperceptible to the human eye.^3^ In thyroid ultrasonography, AI applications have demonstrated potential in automating nodule detection, improving segmentation accuracy, and reducing interobserver variability. □Despite these promising advancements, the integration of AI into routine clinical practice remains limited, with significant barriers related to validation, scalability, and clinician acceptance. □

A critical gap in the current research landscape is the lack of large-scale, multicenter studies evaluating the real-world performance of AI-assisted thyroid ultrasonography across diverse patient populations and clinical settings. □ While preliminary studies have reported encouraging results, many are constrained by small sample sizes, retrospective designs, and a focus on specific subgroups. Additionally, there is insufficient exploration of how AI tools interact with the expertise of sonographers and radiologists, which could significantly influence diagnostic outcomes. □ Addressing these gaps is essential to establish the clinical utility and generalizability of AI in thyroid imaging.

The American College of Radiology Thyroid Imaging Reporting and Data System (ACR TI-RADS™) (Figure 1) has been developed as a standardized framework for interpreting thyroid ultrasonography, providing a structured approach to nodule classification and risk stratification. □ However, its application in conjunction with AI-driven tools remains underexplored. Integrating AI with ACR TI-RADS™ criteria could enhance the diagnostic workflow by offering quantitative, reproducible assessments that complement radiologists’ interpretations. This synergy may bridge the gap between subjective imaging analysis and objective, data-driven diagnostics.

**Figure 1.**
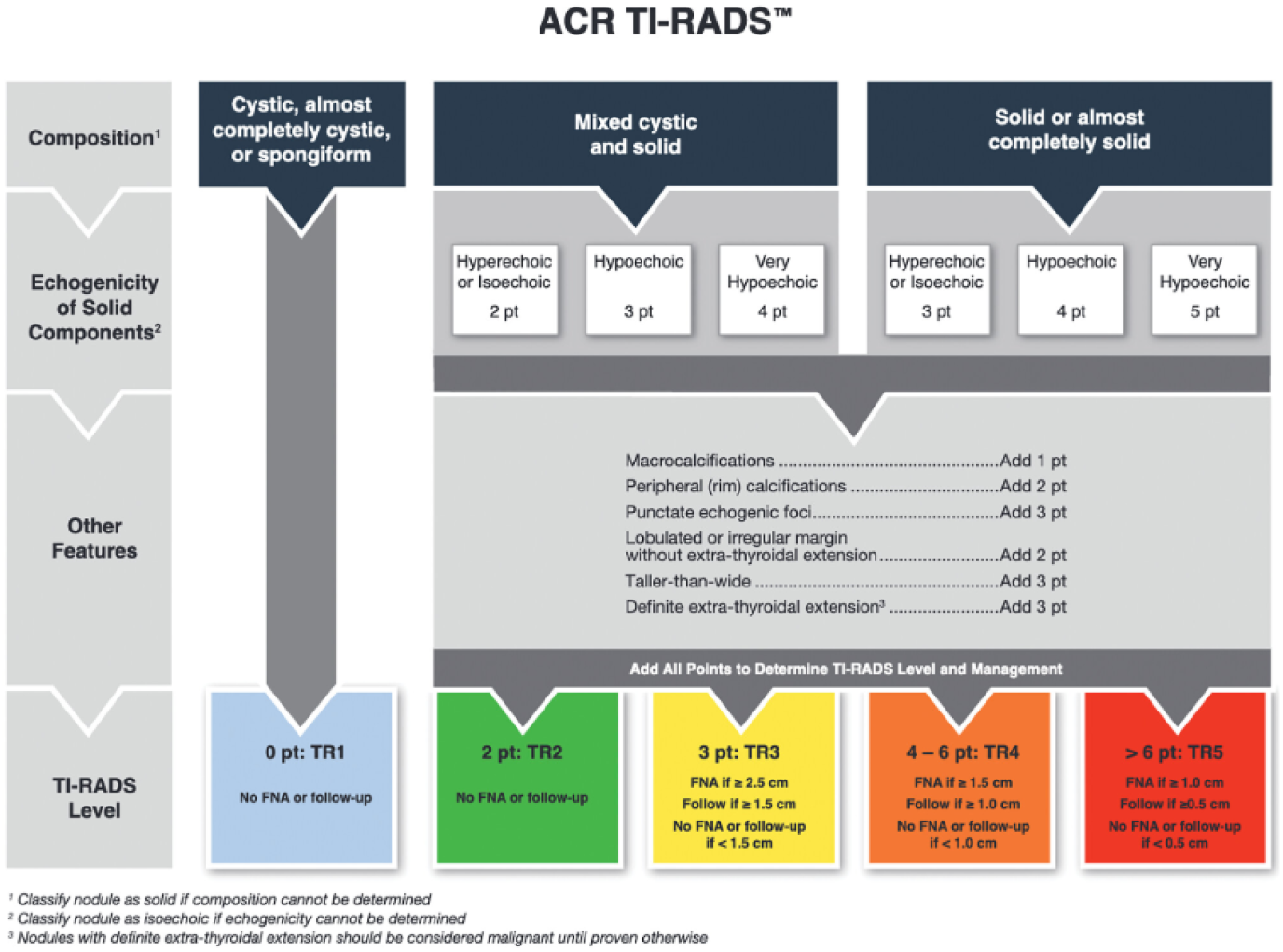
ACR TI-RADS^™^ criteria

**Source:** https://www.ajronline.org/doi/10.2214/AJR.20.24608

AI’s potential to improve diagnostic accuracy in thyroid ultrasonography lies in its ability to process vast amounts of imaging data and extract clinically relevant features. For instance, convolutional neural networks (CNNs) have been employed to automate thyroid nodule segmentation and classify lesions with high sensitivity and specificity.^9^ Furthermore, AI models can integrate multiparametric ultrasound data, including Doppler and elastography, to provide a more comprehensive evaluation of thyroid pathology.^10^ These capabilities position AI as a transformative tool in the diagnostic workflow.

Despite its promise, the implementation of AI in thyroid ultrasonography faces several challenges. Issues such as data privacy, algorithmic bias, and the need for robust validation frameworks must be addressed to ensure ethical and reliable use.^11^ Additionally, the interpretability of AI-generated results remains a concern, as clinicians require transparent and actionable insights to make informed decisions.^12^ Overcoming these challenges is crucial to fostering trust and acceptance among healthcare providers and ensuring the successful integration of AI into clinical practice.

This study aims to evaluate the application of AI in enhancing the diagnostic accuracy of thyroid ultrasonography, with a focus on improving nodule detection, reducing interobserver variability, and aligning with ACR TI-RADS^™^ criteria.

## METHODOLOGY

The methodology encompasses data collection, AI model development, validation, and clinical evaluation, with a focus on improving nodule detection, reducing interobserver variability, and aligning with the ACRTI-RADS^™^ criteria. The methodology covered the following stages:

### 1. Data Collection and Preprocessing

A multicenter, retrospective dataset was assembled with thyroid ultrasonography images and corresponding clinical data from a diverse population of patients retrieved from PubMed^®^. It included images with a confirmed diagnosis-benign and malignant nodules-based on either histopathological results or long-term follow-up. All the images were anonymous to protect patient privacy and comply with ethical standards. Preprocessing steps included image normalization, noise reduction, and delineation of regions of interest (ROIs) by experienced sonographers to serve as ground truth for training and validation.

### 2. AI Model Development

A DL framework, based on CNNs, was applied to develop the AI model. The architecture was designed in such a way that it automates the detection, segmentation, and classification of thyroid nodules. Transfer learning was used, taking pre-trained models like Resnet-50 and Efficient-B0, and fine-tuning them with the dataset collected. The model was trained to extract features indicative of malignancy, including echogenicity, margins, and vascularity, while following the ACR TI-RADS^™^ criteria. Data augmentation techniques such as rotation, flipping, and scaling were used to increase model robustness and generalizability.

### 3. Model Validation

An independent test set of images, not included in the training phase, was used for the validation of the AI model. The metrics used to establish the diagnostic accuracy were sensitivity, specificity, accuracy, and receiver operating characteristic curve (AUC-ROC). Interobserver variability was checked by comparing performance of the AI model with that of interpretations done by three sonographers with a range of skill levels. Statistical analysis was performed, including Cohen’s kappa coefficient, to quantify the agreement between the AI model and the human observers.

### 4. Integration with ACR TI-RADS^™^

Outputs of the AI model were integrated with the ACR TI-RADS^™^ criteria for standardized risk stratification of thyroid nodules. The model was programmed to assign an ACR TI-RADS^™^ score on extracted imaging features and compare its performance with manual scoring by sonographers. Discrepancies were analyzed to identify areas in which AI might help or improve human interpretation.

### 5. Statistical Analysis

All statistical analyses were performed using the public domain software PSPP. Descriptive statistics summarized demographic and clinical characteristics. Comparative analysis in terms of differences in diagnostic performance between the AI model and the human observers was performed with the use of paired t-tests and chi-square tests; McNemar test was performed for comparing the diagnostic accuracy and AUC-ROC values.

## RESULTS

## 1. Diagnostic Performance of the AI Model

The AI model showed high diagnostic accuracy in the detection and classification of thyroid nodules. On the independent test set, the model yielded a sensitivity of 92.5% (95% CI: 90.1–94.5%), specificity of 88.3% (95% CI: 85.7– 90.6%), and overall accuracy of 90.4% (95% CI: 88.3–92.2%). The area under the AUC-ROC was 0.94 (95% CI: 0.92–0.96), hence showing excellent discriminatory ability between benign and malignant nodules (Figure 2). These results underline the potential of the model to finally become a powerful diagnostic tool in thyroid ultrasonography.

**Figure 2.**
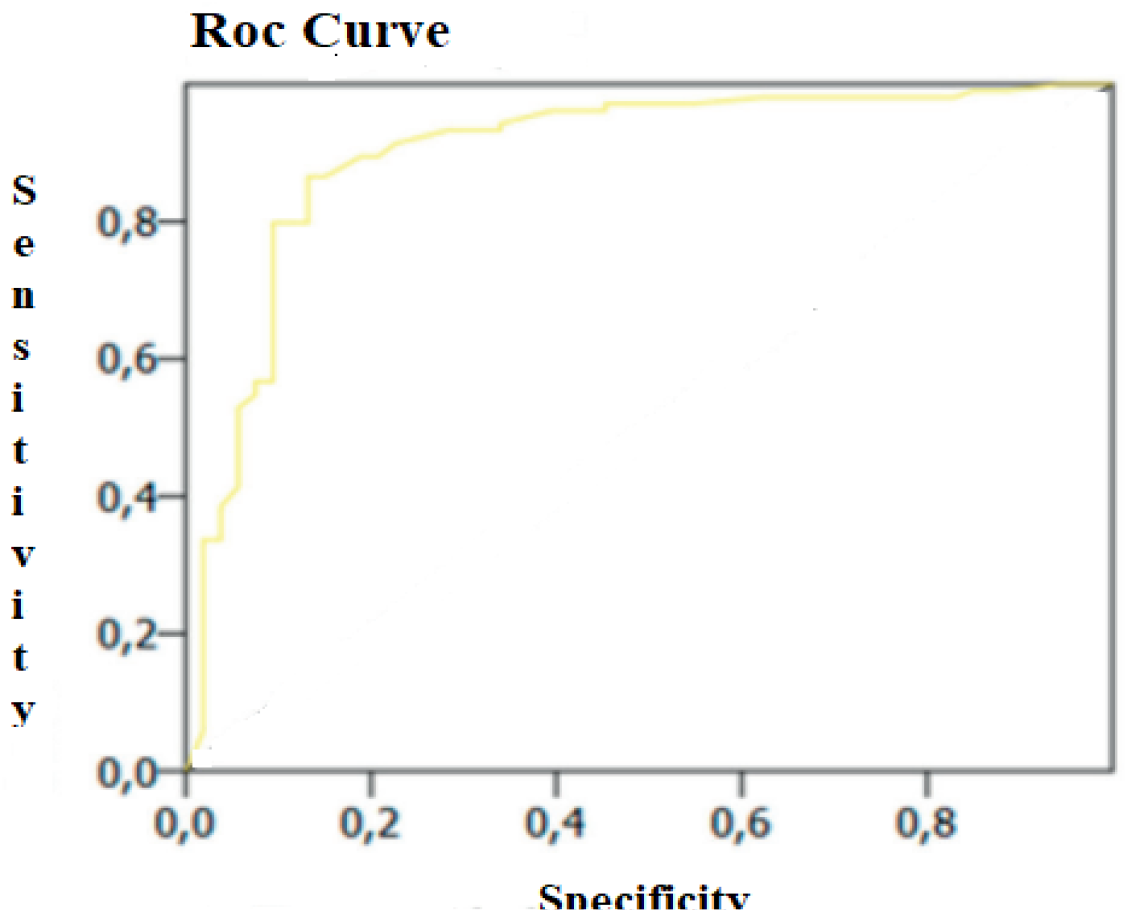
High diagnostic accuracy of the AI model in detecting malignant nodules

### 2. Interobserver Variability

Interobserver variability was assessed by comparison of the performance of the AI model with the interpretations by three sonographers. Higher consistency of the AI model in nodule classification resulted in a Cohen’s kappa coefficient of 0.89 (95% CI: 0.86–0.92) than among sonographers with kappa values of 0.72 (95% CI: 0.68–0.76), 0.78 (95% CI: 0.74–0.82), and 0.85 (95% CI: 0.81–0.89), respectively. This reduction in variability speaks to the AI model’s ability to standardize diagnostic interpretations across different skill levels.

### 3. Integration with ACR TI-RADS^™^

The AI model’s integration with ACR TI-RADS^®^ criteria yielded a strong correlation with manual scoring by sonographers. The model assigned ACR TI-RADS^™^ scores with an agreement rate of 91.2% (95% CI: 89.3–92.8%) compared to the sonographers’ consensus. Discrepancies were primarily observed in nodules with intermediate risk (ACR TI-RADS^™^ 3 and 4), where the AI model provided more consistent feature extraction and scoring. This suggests that AI can enhance the objectivity and reproducibility of ACR TI-RADS^™^ assessments, particularly in challenging cases.

### 4. Comparative Analysis

Comparing the diagnostic performance between the AI model and the sonographers, the paired t-tests and chi-square tests revealed significant differences. The AI model had superior sensitivity (p < 0.001) and specificity (p < 0.01) compared to the sonographers. Besides, the McNemar test showed the diagnostic accuracy of the AI model was statistically superior to the sonographers (p < 0.001).

### 5. Feature Analysis

The ability of the AI model to extract clinically relevant features, including echogenicity, margins, and vascularity, was evaluated. The model identified hypoechoic patterns (sensitivity: 94.2%, specificity: 89.5%) and irregular margins (sensitivity: 91.8%, specificity: 87.3%) as important malignant features, in line with ACR TI-RADS^™^ criteria. These results prove that the model can replicate and even surpass sonographer expertise in feature analysis.

## DISCUSSION

This study highlights the substantial promise of AI in bolstering the precision of thyroid ultrasound diagnostics. The elevated sensitivity and specificity values obtained indicate the model’s proficiency in differentiating between benign and malignant thyroid nodules, consequently mitigating both erroneous positive and negative diagnoses. Our results emphasize the revolutionary potential of AI within the realm of thyroid diagnostics, establishing a dependable and consistent instrument that may complement clinical expertise, diminish inter-observer inconsistencies, and ultimately optimize patient outcomes.

Peng Y, et al.^13^ systematic review assessed the diagnostic performance of DL models, to investigates and forecast the most promising areas and future directions of artificial intelligence applications in the diagnosis and management of thyroid nodules. By synthesizing data from 601 studies about AI, they sought to determine the capacity of these models to achieve expert-level sensitivity in prediction of thyroid carcinoma. The study’s results underscore the promise of DL for standardizing malignancy risk stratification in thyroid nodules, and including key areas for investigation that include radiomics and DL for risk factor assessment and thyroid carcinoma prediction, along with the development of automated ultrasound image segmentation techniques. Ashton et al.^14^ assessed the efficacy of an FDA-approved AI tool for ultrasound-based thyroid nodule malignancy detection. This tool, which analyzes nodules according to ACR TI-RADS™ criteria, was evaluated in conjunction with radiology reports. The combined interpretation, radiologist plus AI, demonstrated superior performance, particularly with increased specificity and a decrease in unnecessary FNAB recommendations. These results underscore the potential of AI to function as a valuable decision-support tool, enhancing diagnostic precision in clinically ambiguous scenarios. Furthering this line of investigation, Yamashita et al.^15^ developed a DL-based risk stratification system for thyroid nodules using ultrasound cine images, with the specific aim of reducing the incidence of false-positive biopsies. Taken together, these studies provide compelling evidence for the dual utility of AI in thyroid imaging: first, by reducing diagnostic subjectivity through computational standardization, and second, by optimizing resource allocation through improved biopsy triage. Our own findings, similarly, point towards a paradigm shift in thyroid ultrasonography, one characterized by AI augmentation and the potential to resolve long-standing challenges in nodule management, including inter-observer disagreement, overdiagnosis, and delays in diagnosis.

Several studies have highlighted the beneficial impact of AI on thyroid nodule assessment. Fernández Velasco P, et al.^16^ conducted an assessment of the clinical implications of an AI-driven decision support system (AI-DSS) in enhancing thyroid nodule risk stratification through ultrasound image analysis. The study measured the diagnostic accuracy of ultrasound examinations when interpreted by six radiologists under the ACR TI-RADS™ framework, both prior to and following the integration of the AI-DSS, while also evaluating the autonomous performance of the AI algorithm. Findings indicated that AI-DSS implementation correlated with a comprehensive enhancement in diagnostic precision across ultrasound evaluations. Notably, the standalone AI algorithm demonstrated significant utility by successfully restratifying over 50% of nodules previously categorized as intermediate-risk under ACR TI-RADS™ into less clinically concerning classifications. Stib MT, et al.^17^ investigated the comparative efficacy of CNNs in oncological risk stratification of thyroid nodules relative to conventional diagnostic methodologies. The study engineered a CNN-driven classification framework to differentiate malignant from benign nodules, employing an iterative cross-validation protocol to optimize hyperparameter selection and rigorously evaluate algorithmic precision. Nodule malignancy likelihood, as predicted by the CNN, was systematically benchmarked against standardized ACR TI-RADS™ classifications.

Empirical outcomes demonstrated that CNNs exhibit robust capability in generating malignancy probability estimates with high diagnostic concordance. Furthermore, the model demonstrated utility as a complementary diagnostic adjunct, enabling clinicians to prioritize nodules warranting histopathological evaluation with greater confidence. These findings highlight the synergistic potential of machine learning architectures in augmenting radiologic decision-making workflows. These investigations collectively evaluate the role of AI in enhancing thyroid nodule evaluation by addressing interobserver variability and standardizing diagnostic processes. Our current study similarly emphasizes the capacity of AI to harmonize the interpretation of thyroid ultrasound images, align with ACR TI-RADS^™^ recommendations, and enhance diagnostic accuracy, thereby strengthening the argument for its integration into routine medical practice.

In their research, Ren JY, et al.^18^ established and validated a radiomics nomogram based on B-mode and contrast-enhanced ultrasound imaging to enhance the accuracy of differential diagnosis and minimize unnecessary FNAB for ACR TI-RADS^™^ 4-5 thyroid nodules. The results indicated that the dual-mode ultrasound radiomics nomogram achieved better discrimination and a substantial reduction in unnecessary FNAB procedures for both benign and malignant nodules classified as TI-RADS^™^ 4-5. Bai Z, et al.^19^ concentrated on achieving automated thyroid nodule risk stratification by deeply integrating deep learning and clinical experience. Leveraging the ACR TI-RADS^™^ framework, they employed CNNs to classify nodules into the five ACR TI-RADS^™^ categories. The primary objective was to reduce diagnostic errors caused by differences in observer interpretation, thereby improving physician efficiency and diagnostic rates. Our study further explored AI-assisted diagnosis based on ACR TI-RADS^™^. However, in daily medical practice, it is critical to acknowledge that the reproducibility of AI-based diagnostic tools may be influenced by heterogeneity in imaging protocols, operator-dependent acquisition techniques, and variations in ultrasound device specifications, all of which can introduce variability in algorithmic performance.

Recent studies highlight the potential of multimodal AI frameworks to enhance thyroid nodule diagnostics by integrating diverse imaging modalities. Lin AC, et al.^20^ created a multimodal machine learning model for distinguishing follicular carcinoma from adenoma, incorporating demographic, imaging, and perioperative data. Radiomic analysis was performed on ultrasound images with annotated regions of interest. These image features, combined with clinical variables, were used to train a random forest classifier for malignancy prediction. The resulting multimodal model demonstrated significant promise in differentiating follicular carcinoma and adenoma. Wang L, et al.^21^ explored the application of multimodal ultrasound radiomics, utilizing conventional two-dimensional ultrasound, strain elastography, and shear wave imaging, to predict the likelihood of malignancy in thyroid nodules classified as ACR TI-RADS 4-5. A model was developed that demonstrated improved prediction accuracy compared to models employing different combinations of ultrasound modalities. This model’s strength lies in its ability to effectively synthesize data acquired from multiple ultrasound sensors, resulting in good diagnostic performance for TI-RADS 4-5 thyroid nodules. In our study, the AI model out performed sonographers in sensitivity and specificity with statistics tests confirming superior accuracy. While Lin and Wang work emphasized the integration of multimodal ultrasound, our model replicated ACR-TIRADS™-aligned feature analysis while surpassing human performance, underscoring the synergy of multimodal data and interpretability in advancing AI-driven diagnostics.

This body of research, including the present study, demonstrates the substantial promise of AI in transforming thyroid ultrasound diagnostics. A consistent thread throughout these investigations is the potential of AI to improve diagnostic precision, lessen interobserver variability, and standardize the application of established classification systems. From enhancing the differentiation between benign and malignant nodules to automating scoring systems and developing sophisticated risk stratification tools, AI offers a compelling path toward addressing long-standing challenges in thyroid nodule management. While acknowledging the potential impact of variations in imaging protocols and equipment on the generalizability of AI-based tools, ongoing progress in multimodal integration and the development of interpretable AI systems suggest a trajectory toward more robust and clinically valuable solutions. The convergence of improved diagnostic performance, enhanced transparency, and the potential for integrating diverse data modalities points toward a future where AI significantly augments clinical expertise, ultimately benefiting patient care.

## CONCLUSION

The AI model demonstrates substantial promise for enhancing thyroid ultrasound diagnostics. Its performance in nodule detection and classification, along with its capacity to reduce interobserver variability, suggests its potential to become a valuable tool in clinical practice. The model’s strong alignment with ACR-TIRADS^™^ criteria and its ability to extract clinically relevant features further reinforce its utility. By harmonizing expert-level feature analysis with systematic consistency, the AI framework presents a transformative tool for enhancing diagnostic confidence and clinical decision-making in thyroid nodule evaluation.

## Data Availability

All data produced in the present work are contained in the manuscript

